# Transmission modeling to infer tuberculosis incidence, prevalence, and mortality in settings with generalized HIV epidemics

**DOI:** 10.1101/2022.10.07.22280817

**Authors:** Peter J. Dodd, Debebe Shaweno, Chu-Chang Ku, Philippe Glaziou, Carel Pretorius, Richard J. Hayes, Peter MacPherson, Ted Cohen, Helen Ayles

## Abstract

Tuberculosis (TB) killed more people globally than any other single pathogen over the past decade. Where surveillance is weak, estimating TB burden estimates uses modeling. In many African countries, increases in HIV prevalence and antiretroviral therapy (ART) have driven dynamic TB epidemics, complicating estimation of burden, trends, and potential intervention impact. We therefore developed a novel age-structured TB transmission model incorporating evolving demographic and HIV/ART effects, and calibrated to TB prevalence and notification data from 12 African countries. We used Bayesian methods to include uncertainty for all TB model parameters, and estimated age-specific annual risks of TB infection (ARTI) and proportion of TB incidence from recent (re)infection (PR). We found ARTI of up to 16.0%/year in adults, but a mean PR across countries of 34%. Rapid reduction of the unacceptably high burden of TB in high HIV prevalence settings will require interventions addressing progression as well as transmission.

## Introduction

Over the past decade, tuberculosis (TB) has killed more people than any other pathogen, averaging more than 1.65 million deaths per year between 2010-2019, of which > 363,000 deaths per year were among people living with HIV (PLHIV).^1^ HIV infection has a large impact on TB in individuals: risks of developing TB increase steeply as CD4 cell counts decline, as does the likelihood of death among those not receiving treatment for TB.^2^ Antiretroviral therapy (ART) for PLHIV increases life-expectancy,^3^ and reduces TB mortality^4^ and incidence^5^ (although not to the level of HIV-uninfected people).^6^ This has led to very dynamic and evolving TB epidemics in settings with generalized HIV epidemics, with rapid increases in TB notifications driven by declines in mean CD4 cell counts among people living with HIV (PLHIV), followed by declines in TB notifications as ART coverage has increased.^7^ Globally in 2020, 8% of TB incidence and 14% of all TB deaths occurred among PLHIV.^1^ In the World Health Organization (WHO) African region, these figures are 24% and 31%, respectively; 22 out of the 30 WHO high TB/HIV countries are in Africa.^1^

Our understanding of the global epidemiology of TB is based on estimates of disease incidence and mortality, notably those generated annually by WHO. In many countries with high TB incidence, the gap between estimated TB incidence and cases detected and reported as starting anti-TB treatment (notifications) is substantial, complicating surveillance and accurate measurement of epidemic trajectories. WHO has supported nationally-representative TB prevalence surveys in priority countries, as these provide a measure of TB burden that is less subject to bias than notification data.^8–10^ Since 1990, 43 national TB prevalence surveys have been undertaken and reported results,^1^ including 16 in Africa.^9^ The large sample sizes required mean TB prevalence surveys are expensive to execute, and they are typically powered to generate a relative precision of only +/−20%.^10^

Combining data from prevalence surveys with notifications data (and potentially vital registration data) to produce estimates of incidence and mortality in high HIV-prevalence settings requires a framework that can incorporate the evolving relationship between between incidence, prevalence, and mortality, the effects of HIV, and which can formally account for the uncertainty and relative informativeness inherent in each source of data. Current approaches to TB/HIV burden estimation use dichotomized states to represent HIV and ART which do not capture their evolving and context-specific relationship with TB risk.

Models of TB transmission calibrated to empirical data provide a potential alternative approach to estimating global, regional and national TB burden, but have not been used to date due to technical challenges including the complexity of TB models (especially TB/HIV models), the large number of uncertain parameters, and limitations in surveillance data.^11^ Due to slow timescales in TB progression, models need to be run over timescales representing decades, and evolving age-structures are likely to be important determinants of TB trends.^12^ HIV is strongly patterned by age and sex, and evolving population CD4 cell count distributions among PLHIV and increasing ART coverage result in a dynamic relationship between HIV and TB increasing the model complexity. Many of the parameters describing the natural history of TB are imprecisely known due to long timescales, rareness of measurable outcomes, and suboptimal diagnostic accuracy.

Here we present a novel transmission model/calibration framework that incorporates the dynamic complexity of HIV-fueled TB epidemics and demographic change. We are able to calibrate this model to surveillance data in a likelihood-based formal Bayesian approach with all parameters considered as uncertain. In applying this framework to the 12 WHO high TB/HIV countries in Africa with a nationally representative TB prevalence survey, we provide a new approach to estimating TB burden over time in these settings, and estimate new age-specific metrics of transmission.

## Results

Below, we present fits of our simplified TB transmission model (see figure 1) to available demographic and epidemiological data, and our inferred estimates of national-level TB incidence and mortality, the proportion of TB incidence in PLHIV, the proportion of TB incidence due to infection within the last two years, and also TB infection risks and age-specific contributions to transmission.

**Figure 1.**
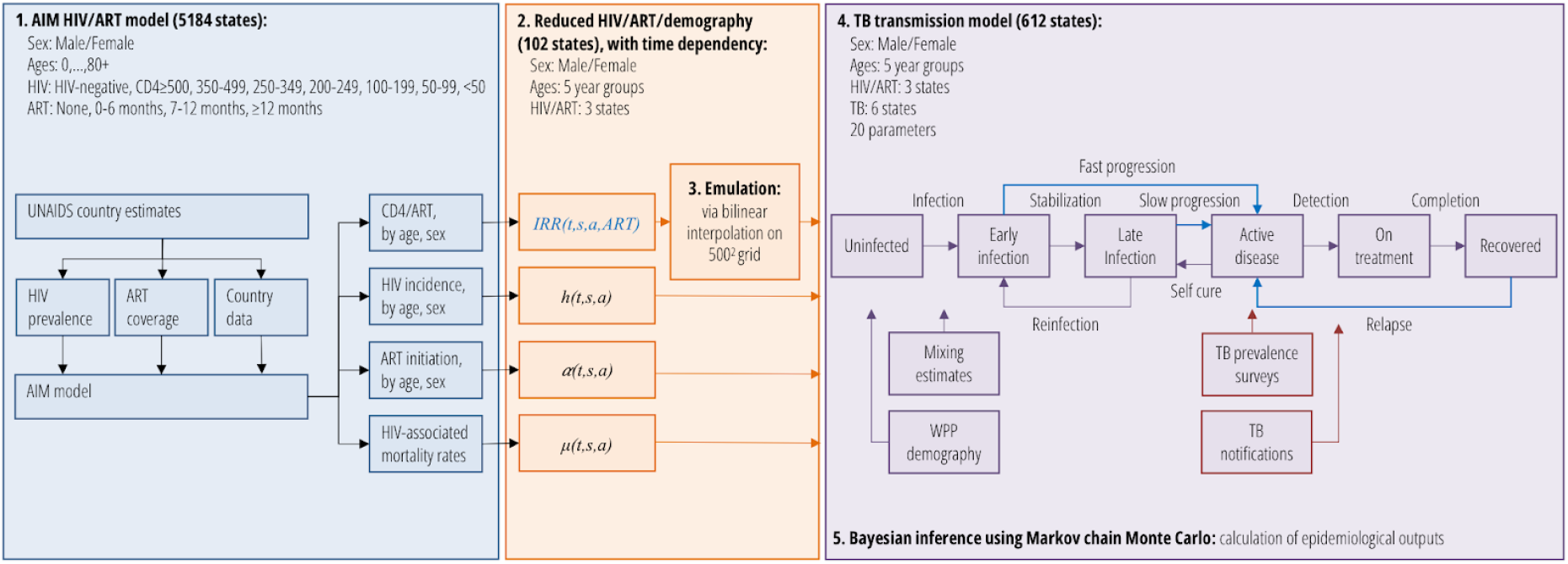
Process flow and model diagram. Blue lines for the TB transmission model in step 4 are processes to which HIV/ART-associated incidence rate ratios *IRR(t,s,a,ART)* are applied. Red boxes in step 4 represent calibration targets.

### Fits to evolving demography and HIV epidemics

Our framework accurately captured changes in national population size and age-sex structure between 1980 and 2019, as well as the number of PLHIV and the number receiving ART (figure 2, rows A and B).

**Figure 2.**
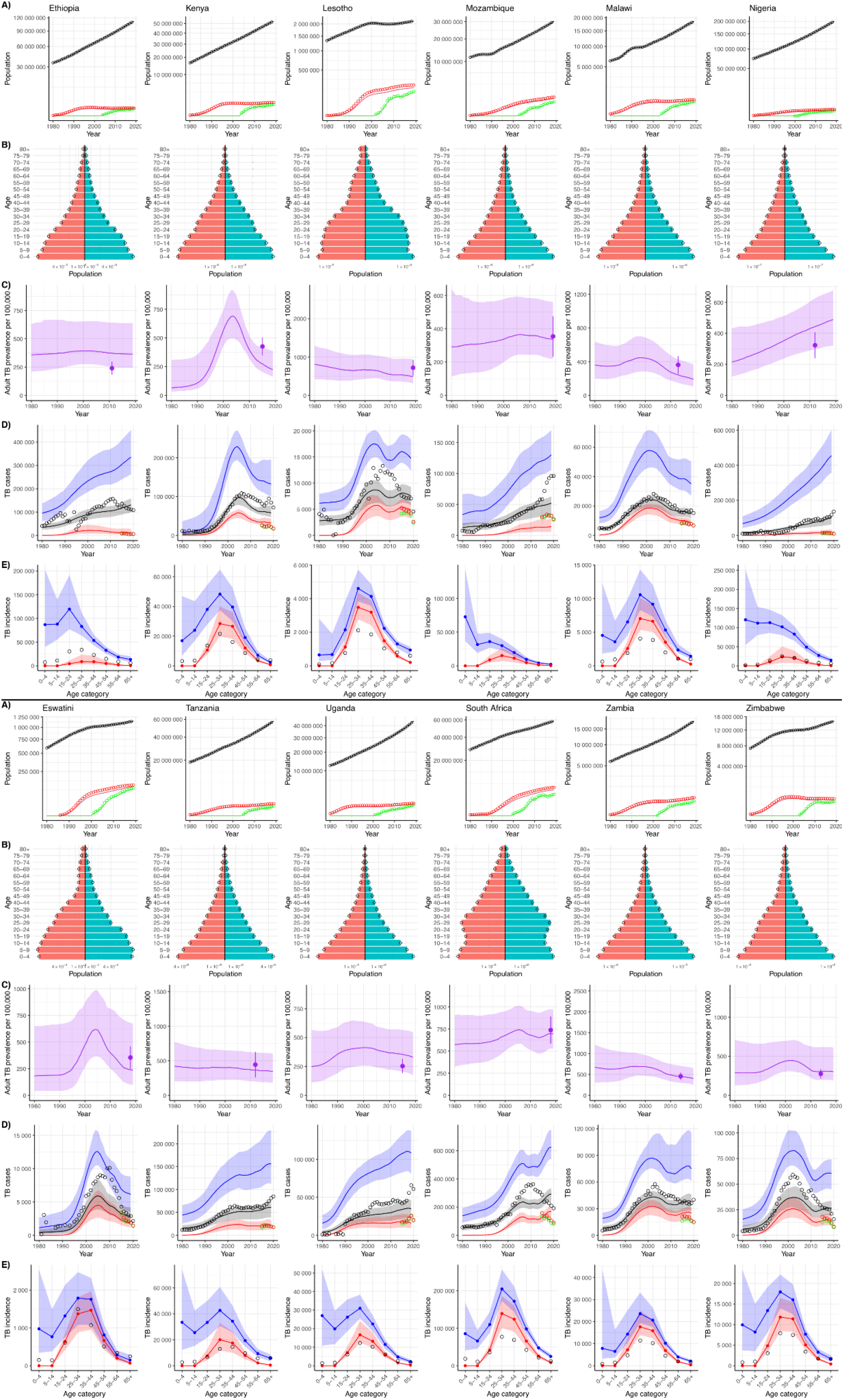
Model outputs compared to empirical data for 12 African countries. Row A) total population (black), people living with HIV (red), people on ART (green) 1980-2019 - points=data, lines=model; Row B) demographic snapshot in 2015 red/left=women, blue/right=men, points=data; Row C) per capita TB prevalence 1980-2019 and 95% credible interval [CrI] as ribbon and TB prevalence data (point/confidence interval); Row D) incident TB 1980-2019, lines=median/ ribbon=95%CrI for model, points=data, blue=TB incidence, black=notified TB, red=notified TB/HIV, green=notified TB/HIV on ART (data); Row E) incident TB in 2015 by age, lines=median/ ribbon=95%CrI for model, points=data, blue=TB incidence, black=notified TB, red=incident TB/HIV.

To make inference tractable, we used a five-year age and sex-structured TB transmission model with HIV/ART (figure 1: all states either HIV-negative, HIV-infected/off-ART, HIV-infected/on-ART, giving a total of 612 states). To capture the dynamics of population immunocompetence, model inputs included results from a modified version of the AIM HIV model (which includes detailed CD4 states and progression, totalling 5184 states) on the time-dependent incidence rate-ratios (IRRs) for incident TB relative to HIV-uninfected, and HIV-related mortality, specific to each age and sex state.^13,14^ Because the parameters governing the link between IRR and CD4 cell-count and describing the protection from ART against TB were included in our inference, we used a bilinear approximation to accurately emulate the parameter dependence of these inputs (see Appendix for results on approximation).

### Fits to TB data

Uncertainty in our estimates of TB-related metrics (figure 2, rows C, D and E) reflected the limited precision of the data and the 20 uncertain parameters around the natural history of TB and case detection, and their interactions with HIV, that were estimated in our Markov chain Monte Carlo (MCMC) inference.

Effective sample sizes (ESS) obtained using an ensemble slice sampling MCMC approach ranged from 393 to 909, with a mean of 605. Priors, posteriors, and bivariate marginal density plots are presented for each country in the Appendix. Judged by means across countries, the strongest five posterior parameter correlations were between: case detection baseline and trend (*cor(K,c)=−0*.*54*); fast progression and transmission (*cor(ε,β)=−0*.*53*); fast progression and IRR CD4-dependence (*cor(ε,α)=−0*.*50*); progression and relative detection in under 5 year olds (*cor(ε*_***04***_,*OR*_***04***_*)=−0*.*33*); and case detection trend and HIV-negative untreated TB duration (*cor(c,D*^*X*^*)=+0*.*30*).

The relative precision of national adult TB prevalence estimates (children aged <15 years are not included in TB prevalence surveys) included in the data likelihood ranged from SD/mean=9% to SD/mean=23% with a median of 13%; the relative precision of our prevalence estimates in years corresponding to prevalence surveys ranged from 16% to 62% with a median of 27%, but was more uncertain in 1980 (figure 2, rows C). Most countries had estimated trends in prevalence that are currently flat or declining, with the exception of Nigeria.

TB incidence estimates (figure 2, row D) were increasing in all countries in the year 2000 due to a combination of population growth and HIV, with some countries experiencing dramatic accelerations.

Per capita incidence has decreased from a peak between 2000 and 2010 in all countries except South Africa and Nigeria (see figure 3), but remains higher in absolute numbers in 2019 than in 2010 for 6 out of 12 countries.

**Figure 3.**
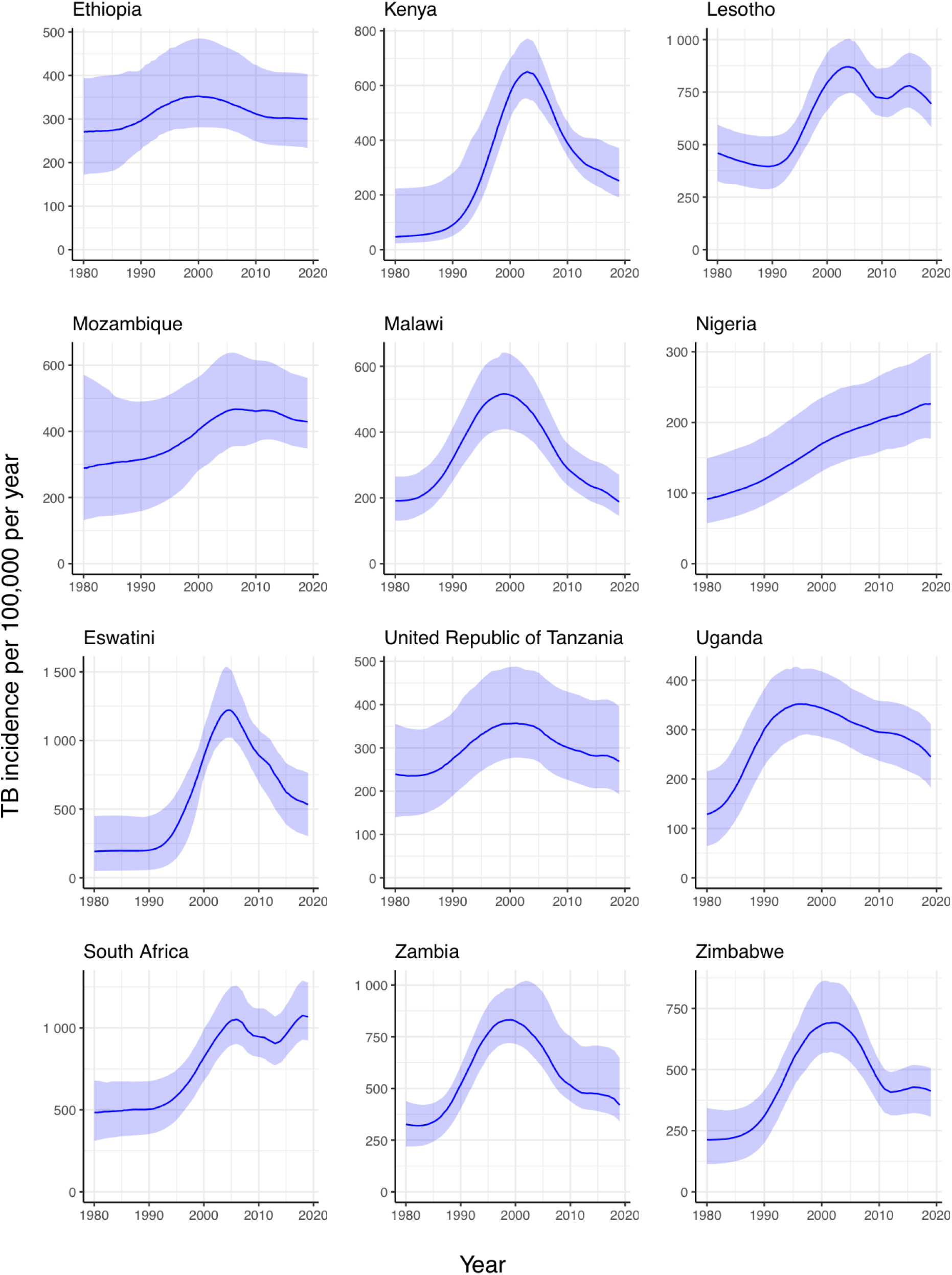
Per capita TB incidence estimated for 12 African countries, 1980-2019. Line=median, ribbon=95% credible interval.

We estimate the TB case detection ratio (the ratio of notifications to incidence in a year) now ranges from 31% (95%UI: 24% to 40%) in Nigeria to 89% (95%UI: 68% to 109%) in Mozambique, with a median over country central estimates of 58%. Case detection was estimated to be lower in younger children. The uncertainty in case detection reflects the weakly informative prior used for the baseline detection probability and the level of noise inferred from the notification data by the hierarchical model.

TB incidence was highest among 25-34 year olds for 9 of the 12 countries (figure 2, row E), a notable exception being Mozambique where age-stratified notifications were absent.

### Comparison with other estimates

There are no data on mortality for direct comparison: among our 12 countries of interest, only South Africa has a vital registration system and substantial coding of HIV deaths as TB means comparison with raw counts is inappropriate. However, our estimates for 2019 were comparable with WHO estimates (figure 4).

**Figure 4.**
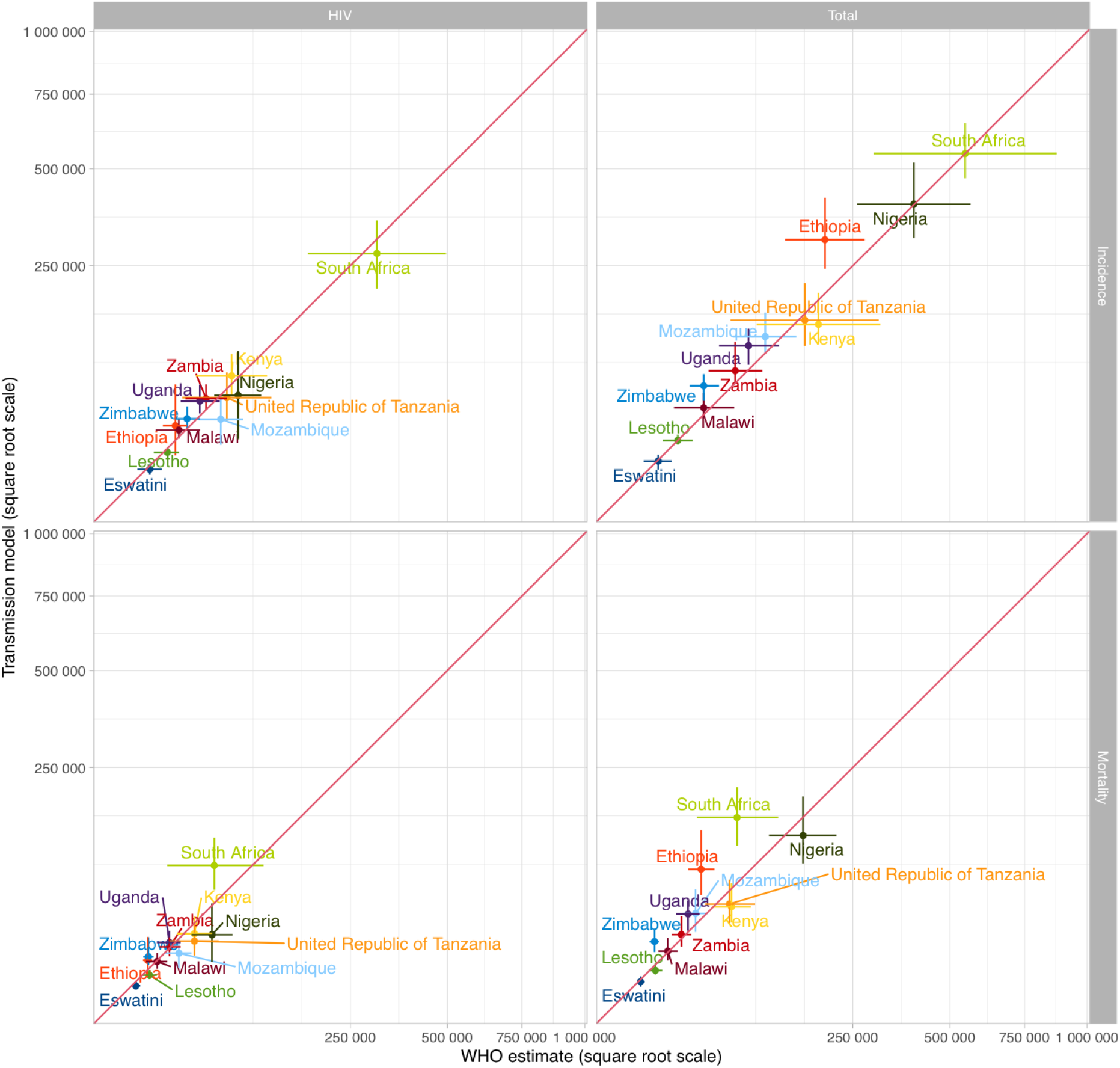
Comparison with WHO incidence and mortality estimates for all TB and TB/HIV for 2019.

### Epidemiological patterns

Our estimates show how the proportion of TB incidence that is HIV associated has evolved over time (figure 5A), reducing in 9/12 countries since 2000, when the mean proportion across countries was 47% to a mean of 41% in 2019. However, this proportion remained high in many countries in 2019, ranging from 7% (95%UI: 2% to 17%) in Ethiopia to 63% (95%UI: 45% to 76%) in Eswatini, despite very high ART coverage.

**Figure 5.**
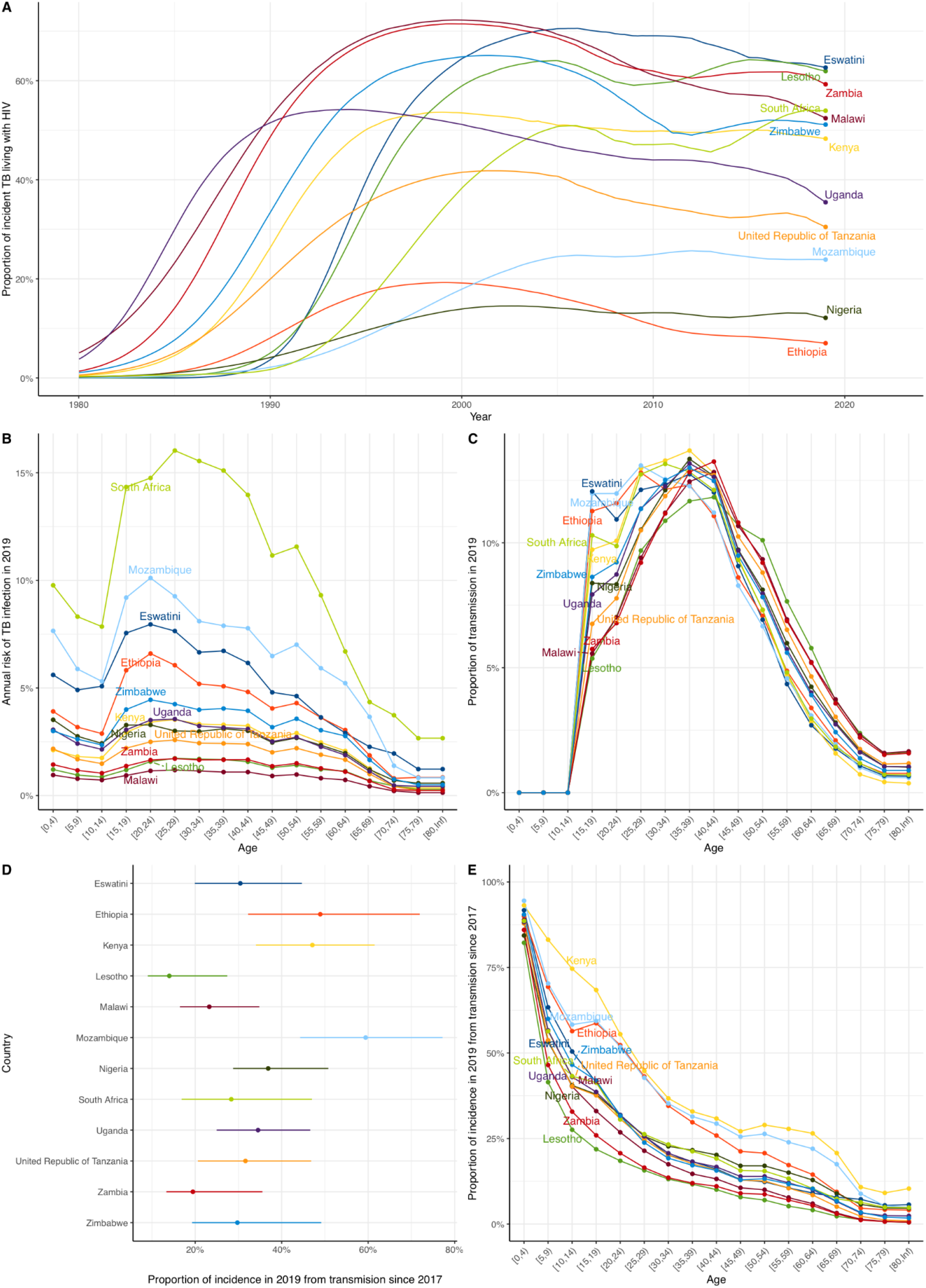
Estimates of other epidemiological metrics. A) The proportion of incident TB that is TB/HIV for 1980-2019; B) annual risk of TB infection in 2019 by age; C) proportion of TB transmission from each age group in 2019; D) proportion of all TB incidence in 2019 due to (re)infection within 2 years; E) proportion of all TB incidence in 2019 due to (re)infection within 2 years for each age group.

We estimate a high and age-dependent annual risk of TB infection (ARTI), reflecting age-assortative mixing and age patterns of infectious TB (figure 5B). The ARTI rose through adolescence typically peaking at 20-30 years of age, with this peak ranging from 1.2%/year (95%UI: 0.7%/year to 2.7%/year) in Malawi to 16.0%/year (95%UI: 7.7%/year to 36.1%/year) in South Africa among 25-29 year olds. Of all TB transmission, we estimate that the highest contribution was from people in the 35-39 year age group, with those aged 25-44 years together accounting for between 44% and 53% of transmission across countries (figure 5C). We found that the mean proportion of TB incidence due to recent infection (within the last two years) across countries was 34% in 2019, and varied from 14% (95%UI: 9% to 27%) in Lesotho to 59% (95%UI: 44% to 77%) in Mozambique (figure 5D). This varied strongly by age, decreasing from a mean over countries of 89% of incidence in those aged 0-4 years to <5% of incidence in those aged over 70 years.

## Discussion

By calibrating a transmission model of TB on an evolving background of demography and HIV-related immunocompetence, we have been able to reproduce patterns in empirical TB data and provide a new method for estimating TB burden in settings with TB prevalence surveys and generalized HIV epidemics. Our model-based account of TB epidemiology suggests strong age-dependent proportions of incidence due to transmission within two years, and that while TB incidence and mortality have declined in the ART era, TB incidence remains enduringly high with nearly half of TB incidence in these countries among people living with HIV, despite rapid scale-up of ART.

Our approach advances the methods used by WHO and IHME for TB burden estimation.^15,16^ In settings with TB prevalence surveys but lacking reliable vital registration data, WHO uses estimates of TB duration and CDR estimates from expert opinion and surveillance system review to estimate incidence and mortality.^15^ IHME estimates are based on a multi-cause regression approach to predicting mortality,^17^ combined with a model of case-fatality and duration (DisMod)^18^ to utilize TB prevalence survey data. Our approach explicitly includes the processes giving rise to TB incidence, prevalence and mortality, includes prior information from our understanding of natural history, and makes only weak prior assumptions about case detection. The level of case detection is determined by the shape of the epidemic and observed prevalence. While our focus here has been on settings where HIV is a key driver of TB, this framework could be easily adapted to lower HIV prevalence settings with TB prevalence surveys. The framework could incorporate vital registration data where available, as well as data not currently used in TB burden estimation, such as TB infection surveys. Application to other settings could quantify the value of information provided by prevalence surveys in terms of more accurate model-based estimates of burden.

TB transmission models at a country level are mainly used to project the impact of interventions, plan health services, and undertake economic evaluations. Probabilistic sensitivity analyses are well-recognized in health economics as an important component of analyses, not just to adequately convey uncertainty to decision makers, but because accurately quantifying uncertainty may influence mean results and therefore the outcome of applying a decision rule.^19^ Fully quantifying uncertainty is more difficult in the case of transmission models because calibration is required to generate parameter samples for analyses. While our emphasis here has been on burden estimation, for which fully capturing uncertainty is also recognized as important,^20^ our technical innovations allowing full characterization of uncertainty based on empirical data would allow economic evaluations of TB interventions to improve their quantification of uncertainty.

A key novelty and strength of this work therefore is in being the first time an age-structured country-level TB transmission model has been calibrated to empirical data using a fully Bayesian paradigm. Almost all country-level TB transmission models are themselves calibrated to modeled estimates, meaning they are not a standalone tool for burden estimation. Similarly, most TB transmission models calibrate considering only a few key ‘free’ parameters as uncertain, meaning their outputs are likely to underestimate uncertainty. Thes practices reflect the technical difficulties of inference with weak data of different types, and the complex, slow and uncertain natural history of TB, implying many parameters with uncertain priors.

Overcoming these challenges has required particular innovations. The first is developing an approach to represent notification and prevalence data in the likelihood which appropriately weights each data source without ad hoc assumptions. We allowed for a natural noise level in the notification data, and by integrating out this parameter prior to inference, we were able to avoid introducing an additional parameter to be sampled. The second is the application of state-of-the-art self-tuning MCMC approaches in order to achieve adequate inference for the 20 parameters in our model. We experimented with a number of tools, mainly developed for cosmological model fitting, and here present results based on the zeus implementation of ensemble slice sampling.^21^ Thirdly, in order to maintain the computational efficiency needed for inference while capturing the full complexity of population immuno-dynamics implied by the AIM model, we used emulation (metamodelling) to capture the changing impact of HIV on TB risk over time, and by age and sex, in our simpler transmission model.

Our work nevertheless has limitations. One important limitation is that while we have included sex strata in our model, we have not modelled sex-specific differences in exposure, natural history, and detection, and therefore not made use of sex-strata in data. In most settings, men have more TB than women, and evidence of poorer care access.^22^ The extent to which this is driven by different exposure, sex-assortative mixing patterns, differences in risk factors or biology, and differences in care seeking is not fully elucidated.^23^ Including sex differences would require several additional uncertain parameters and sex-stratified data to inform them.

Model TB notifications do not fit through the entire history of notification data for all countries. This partly reflects the limited ability of a two-parameter model of case detection to capture any changes over a 40 year period, and in some countries (eg South Africa), implied upticks in TB incidence due to our version of the AIM model slowing down ART initiations so as not to exceed ART coverage data.

We also did not include effects due to COVID-19 and associated changes in mixing and access to care, and for this reason stopped model runs in 2019. COVID-19 typically caused decreases in TB notifications, but the true impact of the COVID-19 pandemic on TB epidemiology is as yet incompletely understood, with potential effects including reductions in community transmission, increases in household transmission, reduced access to care, increased mortality from COVID-19 affecting those at risk of incident TB or with prevalent TB.^24–26^ As understanding of these features improves, these features could in future be included in our framework.

In the countries we considered, the high proportion of TB/HIV on ART emphasizes the importance of more intensive approaches to screening and prevention in these groups. PLHIV taking ART have accessed and remain in regular contact with health services; however, systematic screening for TB and TB preventive therapy should be strengthened. The high estimated ARTI and its pattern by age has implications for the measurement of TB transmission: measures have traditionally been constructed based on surveys in children, but may underestimate (re)infection risks in older age groups, as has previously been suggested.^27,28^

Despite this high ARTI however, we find the proportion of TB incidence due to recent transmission within two years is below half overall in most settings, although higher in younger age groups. Better understanding this metric is important as it quantifies the proportion of incidence that could be averted by interventions to reduce transmission, such as active case finding and improved infection prevention and control.^29^ Clustering metrics from high-resolution molecular epidemiology provide one approach to measurement, although there are few recently published studies from these countries. For Karonga, Malawi, 38% of strains have been reported as genetically linked to other samples within 5 years.^30^ While TB incidence in children and adolescents could therefore respond rapidly to interventions that successfully reduce transmission, such interventions need to be supplemented by strategies that reduce progression from infection to disease (including vaccination), or mitigate the consequences of disease. Our finding that around half of transmission occurs from those aged 25-44 years highlights the importance of effectively engaging working-age adults with health services, including through work-based programmes.

We have demonstrated the potential for calibrated transmission models to be used as a tool for TB burden estimation, and shown it is feasible to calibrate country-level TB models including age and HIV to empirical data while more fully accounting for uncertainty. In doing so, we have developed a number of technical innovations that should be of wider use, especially those working on TB epidemics in high HIV prevalence settings. We provided a rigorous calculation of the proportion of TB incidence due to recent infection, finding substantial contributions from older infections are compatible with high annual TB infection risks in adults. Interventions that reduce TB transmission have the potential to produce rapid and substantial reductions in incidence in younger age groups, but will need to be combined with improved interventions addressing progression to have a major impact on the burden of TB in these settings.

## Data Availability

All data and code are available at https://github.com/petedodd/estevez

## Acknowledgements

This work was funded by an MRC fellowship to PJD (MR/P022081/1) and by the TREATS study (RIA2016S-1632-TREATS); part of the EDCTP2 programme supported by the European Union. This research was funded in whole, or in part, by the Wellcome Trust [206575/Z/17/Z]. For the purpose of open access, the authors have applied a CC BY public copyright licence to any Author Accepted Manuscript version arising from this submission. TC, PM, and PJD were supported by US NIH R01 R01AI147854.

We would like to thank Dr. Rob Glaubius for reviewing an early draft of our description of AIM and clarifying recent changes.

## Methods

### Dynamics of population immunocompetence

We developed a simplified version of the deterministic compartmental AIM model of age- and sex-specific HIV incidence that is used in official UNAIDS estimates.^13,14^ This model includes both sexes (*s*), 81 single-year age groups (*g*), 8 HIV and CD4 cell count categories (HIV-negative, and >=500, 350-499, 250-349, 200-249, 100-199, 50-99, and <50 cells per mm^3^, which we denote *h*),and 4 ART categories among PLHIV (no ART, 0-6 months, 7-12 months, 12+ months, which we denote *r*=0,..,3) for a total of 5184 compartments. We assumed that a TB incidence rate ratio (IRR) at time *t* in each country applied to HIV-related states given by a two-parameter model (ρ controlling the interaction between CD4 decrement and TB risk; *α* controlling the protection from TB due to ART), *irr(t,s,g,h,r*| *α*,ρ*)=<α*^*r/3*^exp[ρΔ_h_]*>*, where <… > denotes an average over CD4 cell decrement below 1000 cells/mm^3^ (Δ_h_) in state *h*; *α* captures the protection from ART in states *r=*0,..,3. We used heuristic pursuit algorithms to achieve fit of this model to the HIV prevalence and ART coverage data used as inputs in the UNAIDS estimates. To facilitate inference, we used the AIM model to calculate mean IRRs and mortality for a model with simpler age (5-year age categories, *a*) and HIV/ART structure (*X = U* for HIV-negative, *H* for HIV-infected/ART-naive, *A* for on ART), for example defining *IRR*^*X*^*(t,s,a*| *α*,ρ*) = <irr(t,s,g,h,r*| *α*,ρ*)>*, where <…> denotes an average over the fine-grained age and HIV/ART states {*g,h,r*} corresponding to the state {*a, X*}. Because parameters *α* and ρ are uncertain and need to vary for inference, we emulated the dependence of these AIM-derived IRRs on *α* and ρ by running the model for a grid of *α* and ρ values and constructing a fast and accurate log-bilinear interpolation approximation to *IRR*^*X*^*(t,s,a*| *α*,ρ) for each country (see Appendix).

### TB transmission model

We used the mean IRRs and HIV-associated mortality computed from this model, together with inputs derived from World Population Prospects demographic estimates^31^ to develop a TB transmission model with the simplified HIV/ART/demographic structure, that reproduced HIV and demographic trends. We used an established TB model structure with 6 states to represent: uninfected, fast-progressing latent infection, slow-progressing latent infection, TB disease, anti-TB treatment, and recovered from previous anti-TB treatment, which together with 17 age categories, both sexes and 3 HIV/ART stages gave 612 ordinary differential equations (see Appendix). Our age-, sex-, and time-dependent TB IRRs were applied to all three pathways for incident TB (recent progression, non-recent reactivation, relapse after successful treatment). We used country-specific estimates of age-dependent mixing rates from Prem et al.^32^ in determining the force-of-infection, and assumed that children under 15 years were not infectious. We developed priors for the parameters of this model and its initial condition in 1970, based on established literature, including different progression rates and durations for children under 5, and odds ratios for detection in the 0-4 year and 5-14 year age categories (see Appendix). The base detection model used a logit-normal prior with mean 0 and standard deviation 0.3 scaled onto the interval [0,0.9) to represent an initial case detection ratio (CDR) for each country, and a linear time trend in logit-space.

### Data and likelihood

To capture the relative influence of TB notification and TB prevalence data, we used a hierarchical model that allowed the noise level for notifications (σ) to be learned from data. To use a single prior for σ across countries, we scaled by the maximum over yearly notifications. We used Gaussian approximations for the prevalence and notification likelihoods. The standard deviation for the prevalence likelihood (σ_*P*_) was based on the empirical precision of the estimate of bacteriologically-positive TB prevalence in those aged 15+ years (*P*_*e*_), so that, dropping constants, the data log-likelihood (*LL*) was given by:

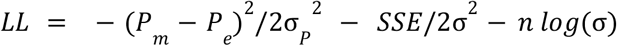

where *P*_*m*_ is the corresponding transmission model estimate of TB prevalence, *n* is the number of years with notification data, and *SSE* is the sum-of-squares difference between TB notifications and corresponding model estimates, divided by the square of the maximum yearly notifications in each country. We chose an uncertain inverse-gamma prior for σ, which enabled us to integrate over σ in closed form, resulting in one fewer parameters to sample from and a marginal likelihood (*ML*):

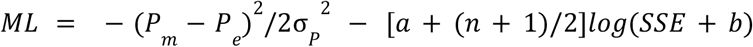

where *a* and *b* are the inverse-gamma hyperparameters (see Appendix). In years where some of the notifications are age-stratified, we split estimates into stratified and unstratified components with a noise term consistent with the total, which results in a modified *SSE* term. While age-specific prevalence estimates are very uncertain, we incorporate this information in the data likelihood using relative rates of prevalence (15-24 years as the reference category) and a Gaussian approximation. Similarly, we calculated a fraction of routine notifications that were among PLHIV and its standard deviation using a random-effects meta-analysis, and used this as a target for the relevant years’ model average corresponding fraction.

We considered the 12 WHO high TB/HIV countries in Africa with a nationally representative TB prevalence survey. Of these, only South Africa had vital registration data. Death coding issues related to stigma mean that deaths recorded as due to TB in South Africa are a substantial overestimate of true TB deaths. We therefore did not include a mortality likelihood component.

### Bayesian inference

After integrating out σ, we were left with 20 parameters whose log prior densities were added to the likelihood. Each country was fitted separately using an ensemble slice sampling MCMC implementation,^21^ using 50 chains and 2,000 iterations.

### Proportion of TB incidence from recent transmission

The proportion of incidence due to recent transmission cannot be reliably calculated as the fraction of incidence from ‘fast’ progressing model compartments, especially when TB/HIV IRRs mean non-negligible fractions of ‘slow’ progressors will develop disease within 2 years. We carefully captured the proportion of incidence due to recent transmission within 2 years in 2019 by calculating incidence in an auxiliary copy of the differential equations with zero initial conditions, populated only by the (re)infection flows in the main model during 2017-2019 (see Appendix).

